# North West London Covid-19 Vaccination Programme: Real-world evidence for Vaccine uptake and effectiveness

**DOI:** 10.1101/2021.04.08.21254580

**Authors:** B Glampson, J Brittain, Amit Kaura, A Mulla, L Mercuri, S Brett, P Aylin, T Sandall, I Goodman, J Redhead, K Saravanakumar, E K Mayer

**Affiliations:** ICT, Imperial College Healthcare NHS Trust, Praed Street, W2 1NY, London, UK; Imperial College London, 10th Floor QEQM building, St Mary’s Hospital Campus Praed Street, W2 1NY, London, UK; Imperial College London and Imperial College Healthcare NHS Trust, Hammersmith Hospital, W12 0HS, London, UK; Imperial College Healthcare NHS Trust, Praed Street, W2 1NY, London, UK; Imperial College London, Hammersmith Hospital, W12 0HS, London, UK; Imperial College London, Medical School Building, St Mary’s Hospital, W2 1PG, London, UK; North West London Collaboration of Clinical Commissioning Groups, 15 Marylebone Road, NW1 5JD, London, UK; Boundary House, Cricket Field Rd, Uxbridge, UB8 1QG, UK; 15 Marylebone Road, NW1 5JD, London, UK

## Abstract

**Objective:** To assess the early vaccine administration coverage and vaccine effectiveness and outcome data across an integrated care system of eight CCGs leveraging a unique population-level care dataset

**Design:** Retrospective cohort study.

**Setting:** Individuals eligible for COVID 19 vaccination in North West London based on linked primary and secondary care data.

**Participants:** 2,183,939 individuals eligible for COVID 19 vaccination

**Results:** During the NWL vaccine programme study time period 5.88% of individuals declined and did not receive a vaccination. Black or black British individuals had the highest rate of declining a vaccine at 16.14% (4,337). There was a strong negative association between deprivation and rate of declining vaccination (r=-0.94, p<0.01) with 13.5% of individuals declining vaccination in the most deprived postcodes compared to 0.98% in the least deprived postcodes.

In the first six days after vaccination 344 of 389587 individuals tested positive for COVID-19 (0.09%). The rate increased to 0.13% (525/389,243) between days 7 and 13, before then gradually falling week on week.

At 28 days post vaccination there was a 74% (HR 0.26 (0.19-0.35)) and 78% (HR 0.22 (0.18-0.27)) reduction in risk of testing positive for COVID-19 for individuals that received the Oxford/Astrazeneca and Pfizer/BioNTech vaccines respectively, when compared with unvaccinated individuals.

After vaccination very low rates of hospital admission were seen in individuals testing positive for COVID-19 (0.01% of all patients vaccinated).

**Conclusions:** This study provides further evidence that a single dose of either the Pfizer/BioNTech vaccine or the Oxford/Astrazeneca vaccine is effective at reducing the risk of testing positive for COVID-19 up to 60 days across all adult age groups, ethnic groups, and risk categories in an urban UK population. There was no difference in effectiveness up to 28 days between the Oxford/Astrazeneca and Pfizer/BioNtech vaccines.

In those declining vaccination higher rates were seen in those living in the most deprived areas and in Black and Black British groups.

There was no definitive evidence to suggest COVID-19 was transmitted as a result of vaccination hubs during vaccine the administration roll-out in NWL, and the risk of contracting COVID-19 and/or becoming hospitalised after vaccination has been demonstrated to be very low in the vaccinated population.

## Introduction

On the 11^th^ of March 2020 the World Health Organisation (WHO) declared the novel coronavirus, referred to as SARS-CoV-2 causing COVID-19 syndrome, as a pandemic with governments across the world implementing restrictive measures to slow the spread of the virus and prompting an international effort to develop an effective vaccine (1).

On the 2^nd^ December 2020 the United Kingdom (UK) became the first country to approve a COVID-19 vaccine, after regulators granted emergency-use authorization to BNT162b2 mRNA (Pfizer and BioNTech) following the publication of results of phase 3 trials (2,3). The UK mass vaccination programme commenced on the 8^th^ December 2020 (2). By the 30^th^ December 2020 the ChAdOx1 nCoV-19 adenoviral vaccine, developed by Oxford University/AstraZeneca, was granted regulatory approval by the Medicines and Healthcare products Regulatory Agency (MHRA) and its use included in the UK vaccination programme (2,4). The Moderna vaccine was the third COVID-19 vaccine to be approved for use by the MHRA on the 8^th^ of January 2021 and further vaccines are in development awaiting approval for use (1). The Joint Committee on Vaccination and Immunisation (JCVI) has established the strategy, on behalf of the Government, for the rapid distribution of a first dose of a vaccine to groups of the population deemed to be most vulnerable to severe infection of COVID-19 (5). By the 26th February 2021 29% of the UK population had received at least one dose of an approved COVID-19 vaccine (6). The JCVI stated target was to have offered a first vaccine dose to everyone in Priority groups 1,2,3, and 4 by the 15^th^ February 2021 (7).

Anticipated vaccination coverage of priority groups has been reduced by vaccine hesitancy which is present in UK and Continental European populations alike (8, 9). To ensure the sufficient and rapid uptake of the offered vaccination programme, identifying and addressing vaccination hesitancy and resistance (i.e., the positions where one is unsure about taking a vaccine or where one is absolutely against taking a vaccine) is essential (10). The use of vaccination centres has been reported to increase vaccine hesitancy, possibly due to fear of transmission, but is the only feasible way of administering large numbers of vaccinations rapidly given logistical and cold storage constraints (11). Identifying and understanding COVID-19 vaccine hesitancy within distinct populations may aid future public health messaging.

Real world data supporting the effectiveness of the vaccination strategy in the UK population is needed to guide health policy. This real-word data-driven evidence study of the UK COVID-19 Vaccination Programme in the Northwest London (NWL) population used a unique dataset established as part of the Gold Command Covid-19 response in NWL (iCARE https://imperialbrc.nihr.ac.uk/facilities/icare/), which included the pre-established Whole System Integrated Care (WSIC) data collated for the purposes of population health in the sector.

WSIC is an innovative data sharing initiative by the NWL Collaboration of Clinical Commissioning Groups (CCGs) and has been designed to improve data sharing and interoperability (WSIC https://www.nwlondonccgs.nhs.uk/professionals/whole-systems-integrated-care-wsic-dashboards-and-information-sharingWSICWebpage). WSIC dashboards link provider data from four acute, two mental health and two community Trusts across eight CCGs, social care data from eight boroughs and 360 GP practices to generate an integrated care record for direct patient benefit. The Covid-19 dashboard allows access to data on vaccination and COVID-19 testing within minutes/hours of the data being recorded in source data systems. The vaccination dashboard utilises general practice (GP) clinical systems (SystemOne, eMIS), pathology laboratories (NWL Pathology and TDL), national COVID test results and daily COVID-19 sitreps from the Northwest London secondary care organisations.

### Aims and objectives

The aim of this study was to assess the early vaccine administration coverage and vaccine effectiveness and outcome data across an integrated care system of eight CCGs leveraging a unique population-level care dataset.

### Study objectives were

1. To describe vaccination coverage across NWL CCGs and identify subgroups according to socio-demographic factors and including where vaccination offer was declined.
2. To investigate the impact of vaccine administration on possible virus transmission by assessing rates of positive testing after vaccination; and examine the potential importance of continued isolation following the delivery of a single dose of a COVID-19 Vaccine.
3. To assess the early effectiveness of Covid-19 vaccination over a ten-week follow-up period stratified across population sub-groups and by vaccine type and compared with rates of COVID-19 positive testing rates in the non-vaccinated population.

## Method

### Study design

The study was a retrospective cohort design. Data were captured to support the NWL response to the COVID-19 pandemic on behalf of NWL Gold Command as part of Whole Systems Integrated Care (22). Anonymised data covering vaccinated and unvaccinated individuals from NWL were accessed in the iCARE system for analysis.

### Participants and setting

All adults over the age of 16, eligible to be offered a COVID-19 vaccine, and registered with a GP, or with a resident postcode, in the NWL catchment area were included in the analysis. The eligible population was considered as a static group over the study period based on data available on 24th Feb 2021.

Vaccinated individuals were defined as persons receiving a vaccine within the NWL vaccine programme time period, considered 8th December 2020 to 15th February inclusive. Vaccination status was provided either directly via acute hubs or via GP Electronic Patient Record (EPR) systems via primary care hubs. The unvaccinated group were considered those that had not received a vaccine during the same NWL vaccine programme time period.

Individuals were counted as declining a vaccine if they indicated that they did not want a vaccine, to their GP, and did not then receive a vaccine. Rates of declining vaccination were calculated using the denominator of those who received a vaccine or those that declined a vaccine. Individuals who initially declined vaccination but then were vaccinated after 15th Feb 2021 and before the 24th Feb 2021 were not included as vaccinated.

Follow up analysis included data until 24th February (inclusive) for both groups, allowing over a week of follow up for all individuals.

### Variables

Data variables were age, gender, ethnicity, contraindications to vaccination, vaccine declined, decile of deprivation (as defined by IMD deciles), care home resident, and housebound status. These data were recorded in GP primary care systems.

Risk groups were defined in WSIC (based on the Joint Committee on Vaccination and Immunisation (JCVI) priority cohorts), these were based primarily on individuals in care homes, then those classed as clinically extremely vulnerable, and then on age groups of individuals. Therefore, in the analysis where risk groups were used it should be assumed that the care home and clinical extremely vulnerable can be of any age. Those in care homes were predominantly, although not exclusively, older individuals. Frontline key worker status could not be identified from the data available and therefore could not be analysed separately.

Outcomes measured were the date of result for the first positive swab for all individuals (lateral flow test results were excluded), results included tests from pillar one and two (2). All non-positive (negative, inconclusive and error) were grouped as non-positive results, with the assumption that all non-negative tests would be followed with a second test and these positive results would be included if returned.

Follow-up started on 8th December 2020 for persons not vaccinated and the day of vaccination for those who were vaccinated. Individuals were followed up until outcome measurement or censoring on 24th February 2021.

The denominator for the week-on-week population groups was calculated based on the number of individuals with follow-up data available up to the start of each weekly time period and who had not previously tested positive.

Secondary outcomes of hospitalisation due to COVID were measured as patients who first tested positive after vaccination and prior to or up to seven days after admission (based on PHE guidance) (12), all secondary care data were recorded from patient-level situation reports (sitreps) data submitted by NWL secondary care organisations. This does not include diagnosis data or reason for admission to hospital.

Individuals that received Moderna vaccines (n=3) were excluded from analysis comparing vaccination types due to low numbers.

Patients who died (all cause) between the 8th December 2020 and 24th February 2021 were excluded from the main analysis and included in a sub-analysis, as date of death in the upstream systems is updated variably and therefore likely to be an underestimate.

### Data sources

Whole system integrated care (WSIC) systems data were combined to create an analysis database including (a) GP EPR systems (COVID test results (pillar 2), Vaccination status, type, contraindications, declined, age, gender, ethic group, clinically extremely vulnerable), (b) social care data (care home/housebound), (c) pathology labs (COVID test results (pillar 1)) and NWL acute Trust sitreps (admissions, discharge).

### Identification of Bias

Variations in prevalence of COVID-19 in the population across the timescale of this longitudinal study may alter the rate of positive testing in both the vaccinated and unvaccinated groups. To address these potential confounding factors, prevalence of positivity in the background population and the rate of vaccination delivery were compared.

Unequal use of vaccine type across risk cohorts could make a direct comparison of vaccine outcome data unreliable. We have stated the delivery rates of vaccination types and adjusted denominators appropriately for return to follow up.

Individuals with COVID that did not test positive (untested or asymptomatic) would be included in the COVID negative population. It was assumed that individuals not testing positive were negative. The dataset does not include lateral flow positive tests, which may be more represented in key frontline workers, although frontline workers make up a minority number of the overall NWL population.

The cause of admission of patients was not provided in the acute sitreps and therefore was not available. It was assumed that a COVID-related admission would include any patient testing positive in the period prior to an admission or within seven days of an admission, as per the PHE definition. It was not appropriate to compare hospital admissions between vaccinated and unvaccinated groups as the vaccination programme has targeted the most high-risk individuals, with therefore a presumed higher risk of admission, due to comorbidities.

### Statistical Methods

Multivariable Cox regression analysis was applied to investigate whether vaccination independently predicted having a COVID positive swab during follow-up, after adjusting for age, gender, ethnicity, index of multiple deprivation (IMD) and vaccination manufacturer. Cumulative COVID positive results were graphically displayed accounting for censoring time.

Known missing data included vaccination type for <1% of vaccinated individuals, these data were included in analysis of overall vaccinations, but excluded from vaccination type breakdowns (unless indicated).

Pearson Product-Moment Correlations were used to measure correlations between continuous variables.

### Ethics

This study was approved by NWL Data & Analytics Gold Command and used data collected as part of routine care delivery in NWL London by care providers and data controllers. Data are collated for direct care and population health purposes as part of Whole Systems Integrated Care. All data utilised in the research paper was fully anonymised before analysis and permission was granted to extract aggregate results from the iCARE system.

## Results

### Vaccination Coverage

In NWL, 2,183,939 individuals were eligible to receive a COVID-19 vaccine. 1,059,280 (48.5%) were female, 930,877 (42.6%) were White, 529,492 (24.2%) Asian or Asian British, 166,011 (7.6%) Black and Black British, 60,483 (2.8%) Mixed Race, 189,877 (8.7%) other ethnic group. There was no ethnicity recorded for 307,099 (14.1%).

The week on week testing rate as a proportion of the overall NWL eligible population reached a peak of 1.39% of the population by the week commencing 5th January 2021. After this, it fell to 0.73% of the population in the week commencing 9th February 2021. Eligible population prevalence of positive cases in a week peaked in early January at 0.32% and then fell steadily each week to 0.06% of the population in the week commencing 9th February 2021, with the average across all weeks in the study being 0.19% (Figure 1).

**Figure 1:**
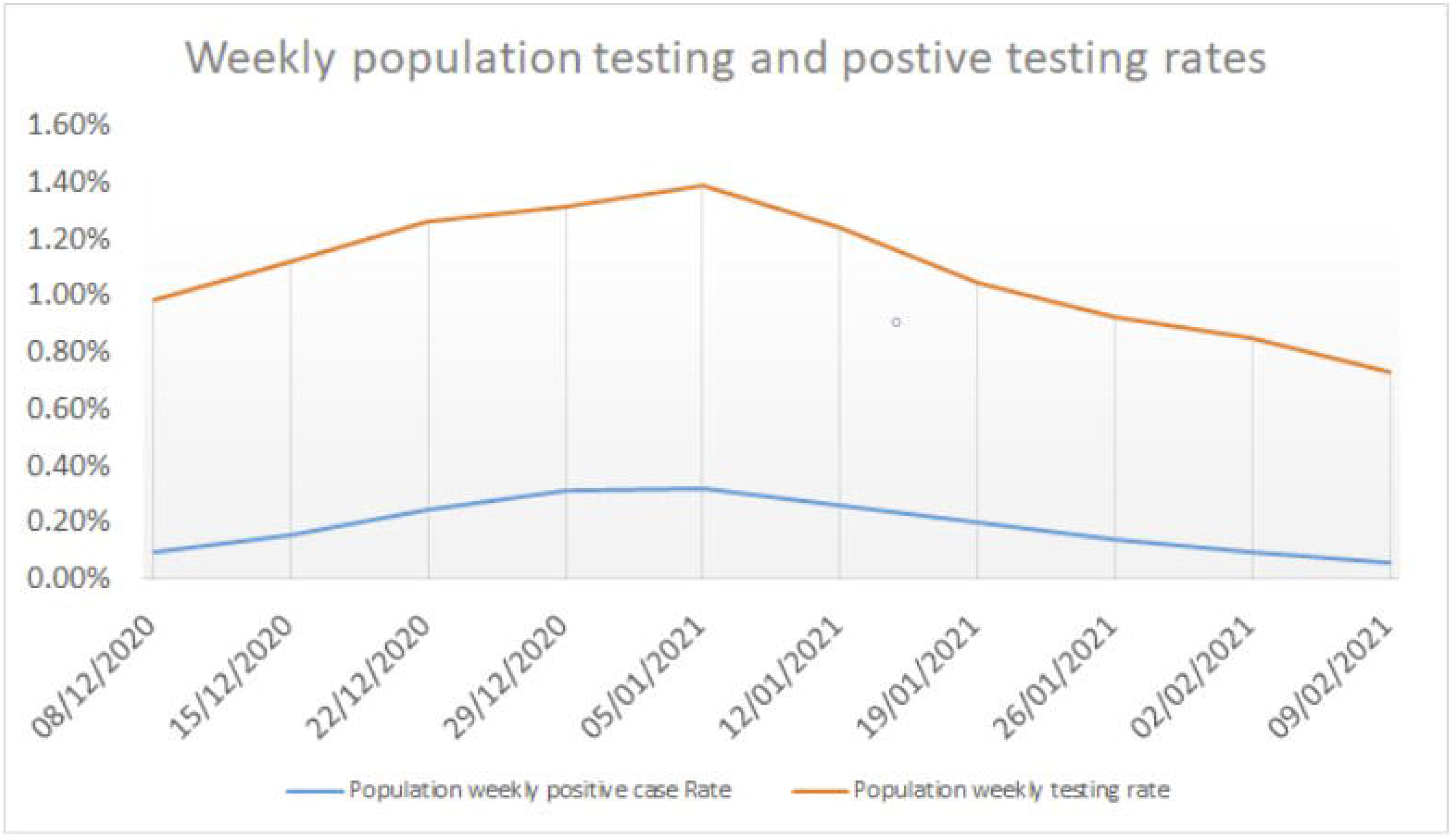
Population weekly person testing rate compared to overall weekly positive case rate in population eligible for vaccination

By 15th February 2021, 389,587 (17.84%) individuals had received at least one dose of a COVID-19 vaccine. Vaccination administration notably increased from early January 2021 with the period between 5th January and 15th February 2021 accounting for 363,304 (93.25%) of the total 389,587 vaccines administered (Figure 2). The number of Oxford/AstraZeneca vaccines administered started to reach parity with Pfizer/BioNTech by Mid-January. In the NWL vaccination programme time period overall, 223,201 (57.29%) Pfizer/ BioNTech and 163,452 (41.96%) Oxford/AstraZeneca vaccines were administered.

**Figure 2:**
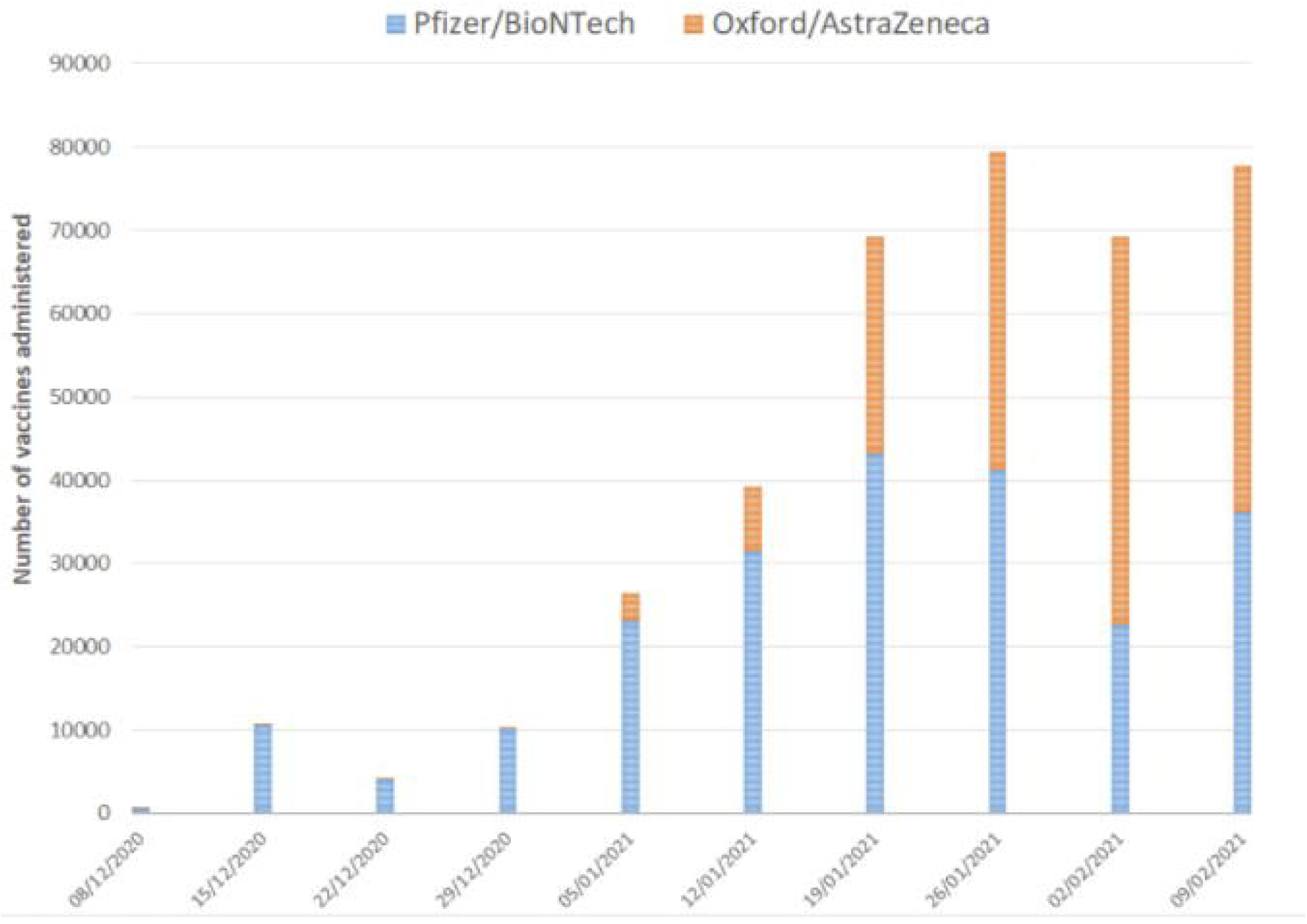
Weekly vaccination numbers of first dose vaccinations given per week from the 8/12/2020 (numbers of vaccines administered defined as all vaccines delivered within the seven-day period from the weekly start date indicated).

During the NWL vaccine programme time period 413,919 individuals were offered a vaccine and 24,332 (5.88%) people declined and did not receive a vaccination. In the vaccinated group 2,957 patients had initially declined but subsequently went on to receive a vaccination; indicating a hesitancy rate of 0.71% (where an individual is initially unsure about taking a vaccine).

Overall, 41.96% of individuals received the AstraZeneca vaccine and 57.29% Pfizer. Pfizer was administered in the majority of individuals aged 16-49 (66.80% n=47817), 75-79 (61.74% n=25348) and 80+ (72.42% n=42090). In those aged 50-74, Pfizer and AstraZeneca were administered with similar proportions (Pfizer - 51.3% n=89,419) AstraZeneca was administered to the majority of care home residents (73.70% n=3822) and in the clinically extremely vulnerable (54.5%, n=21014).

The rate of declining a vaccination across all black, Asian, and minority ethnic groups was 6.39% (11,528) compared with the white group at 4.92% (9,788). Black or black British individuals had the highest rate of declining a vaccine at 16.14% (4,337). Mixed ethnicity groups vaccine declining rate was 10.39% (895). In the Asian and Asian British groups, the rate of declining vaccines was the lowest at 3.21% (3,867). Other ethnic groups declination rate was 9.95% (2,429) and the ethnicity unrecorded group declination rate was 8.52% (3,016). Within the Black or black British individuals, the highest rates of declining vaccination were seen in those aged 80+ or clinically extremely vulnerable at 27.58% and 23.97% respectively (Figure 3).

**Figure 3:**
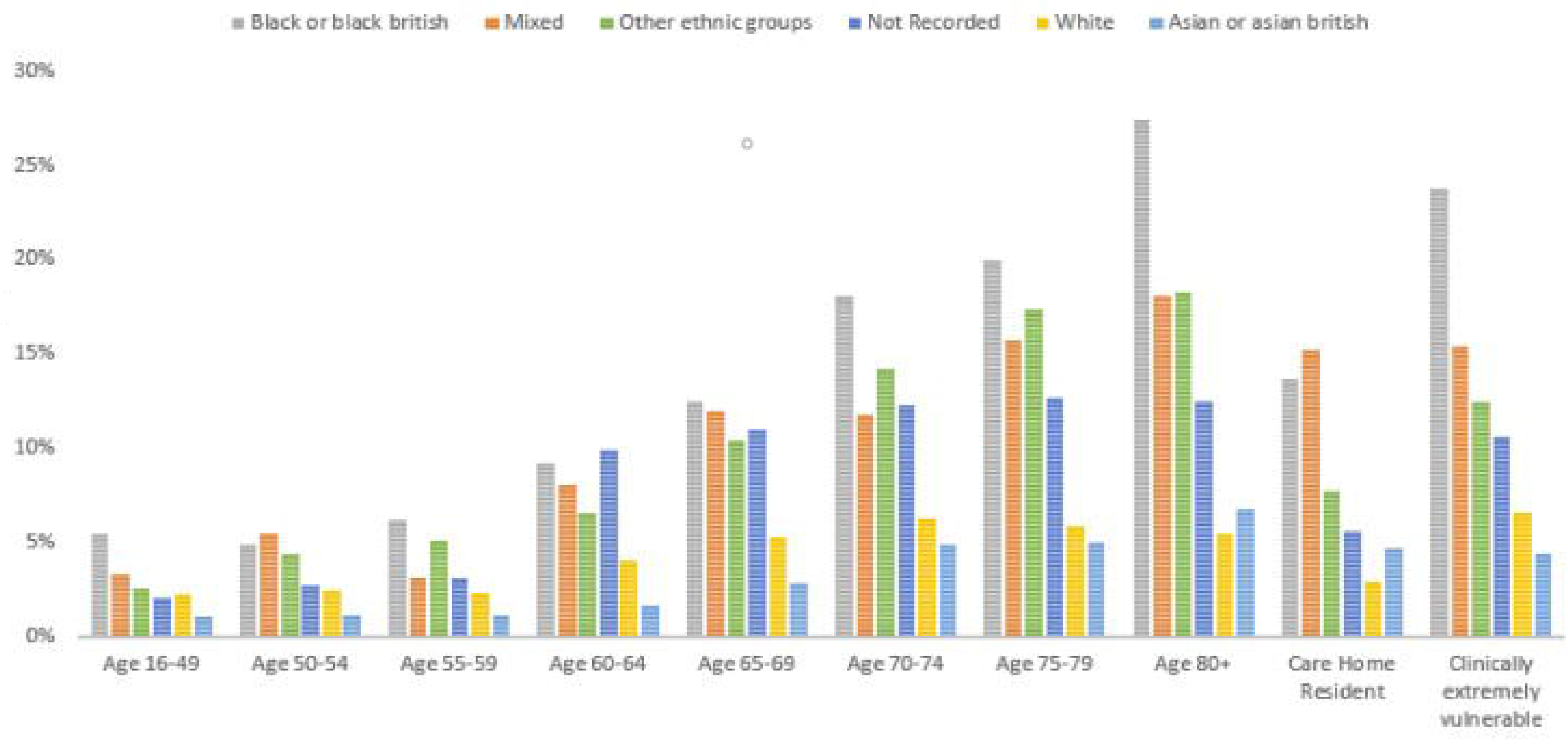
The rates of declining vaccination across WSIC risk categories according to ethnicity

Overall, there were no differences in rates of declining vaccination between gender (female = 5.91%, male = 5.83%). Younger males were more likely to decline than younger females (aged < 65, female = 2.17%, male = 3.16%), whereas older females were more likely to decline (aged 65+, female =7.94%, male = 7.08%).

There was a strong negative association between deprivation and rate of declining vaccination (r=-0.94, p<0.01) with 13.5% of individuals declining vaccination in the most deprived postcodes compared to 0.98% in the least deprived postcodes.

For individuals living in the most deprived areas (bottom decile) those with the highest rates of vaccine decline were the over 70-year-olds (70-74 = 17.52%; 75-80 = 18.99%; 80+ = 25.91%), the clinically extremely vulnerable (19.17%) and black and black British (19.97%) communities.

### Impact of vaccine administration on possible virus transmission

In the first six days after vaccination 344 of 389587 individuals tested positive for COVID-19 (0.09%). The rate increased to 0.13% (525/389,243) between days 7 and 13, before then gradually falling week on week (Table 1). By week seven, fewer than 20 persons were testing positive each week (weekly rate <= 0.05% week five onwards). Over the same time period, no appreciable decrease in the amount of testing of the vaccinated population was observed.

**Table 1:** Absolute numbers of first positive tests and rates of testing based on individuals available for follow up (excluding previously positive). Rates are broken down by individuals in care homes or housebound and those in the rest of the vaccinated population. (*low numbers (1–4) have been replaced with <5).

|  | Days after vaccination |  |  |  |  |  |  |  |  |  |  |
| --- | --- | --- | --- | --- | --- | --- | --- | --- | --- | --- | --- |
|  | Week 1<br><7 | Week 2<br>7-13 | Week 3<br>14-20 | Week 4<br>21-27 | Week 5<br>28-34 | Week 6<br>35-41 | Week 7<br>42-48 | Week 8<br>49-55 | Week 9<br>56-62 | Week 10<br>63-69 | Week 11+<br>70+ |
| Vaccinated individuals time to first positive test after vaccination | 344 | 525 | 332 | 147 | 87 | 48 | 16 | 13 | 11 | <5 | 0 |
| Total vaccinated population completed to period of follow up (excluding previously positive patients) | 389,587 | 389,243 | 330,523 | 261,447 | 184,847 | 111,555 | 62,283 | 31,757 | 20,097 | 14,200 | 2,519 |
| Percent first positive individuals by population completed to follow up time to first positive (not previously positive) | 0.09% | 0.13% | 0.10% | 0.06% | 0.05% | 0.04% | 0.03% | 0.04% | 0.05% | 0.01% | 0.00% |
| Vaccinated individuals (excluding care home or Housebound residents) time to first positive test after vaccination | 319 | 470 | 284 | 129 | 71 | 46 | 13 | 10 | 9 | <5 | 0 |
| Total vaccinated population (excluding care home and housebound residents ) completed to period of follow up (excluding previously positive patients) | 373,820 | 373,501 | 315,666 | 248,136 | 173,336 | 105,834 | 59,574 | 30,674 | 19,338 | 13,763 | 2,431 |
| Percent first positive individuals by population completed to follow up time to first positive (not previously positive) (exc. Care home and housebound) | 0.09% | 0.13% | 0.09% | 0.05% | 0.04% | 0.04% | 0.02% | 0.03% | 0.05% | 0.01% | 0.00% |
| Vaccinated care home or housebound individuals time to first positive test after vaccination | 25 | 55 | 48 | 18 | 16 | <5 | <5 | <5 | <5 | 0 | 0 |
| Total vaccinated care home or housebound population completed to period of follow up (excluding previously positive patients) | 15,767 | 15,742 | 14,860 | 13,317 | 11,556 | 5,770 | 2,749 | 1,090 | 771 | 443 | 92 |
| Percent first positive care home or housebound individuals by population completed to follow up time to first positive (not previously positive) | 0.16% | 0.35% | 0.32% | 0.14% | 0.14% | 0.03% | 0.11% | 0.28% | 0.26% | 0.00% | 0.00% |

Care home residents and/or housebound individuals had a higher rate of positivity in the second week post vaccination at 0.35% compared with the non-care home/Housebound group at 0.13% (Table 1). After the second week, the rate of positivity decreased, although it took until week six to reach less than 0.1%. There was a trend to suggest the rate of positivity decrease week on week was slower when compared with the non-care home/housebound group, but absolute numbers of positive cases in care homes/housebound individuals were very low. Overall, the mean age of care home and house bound residents was 80.6 years.

Testing rate was lowest in the 3-to-4-day period either side of the day of vaccination (Figure 4). After vaccination, the testing rate increased and remained on average higher until day 60. Data after day 60 was not included at the daily level due to low numbers. The reduction in positive test results after vaccination could not be attributed to overall reduction in testing over time.

**Figure 4:**
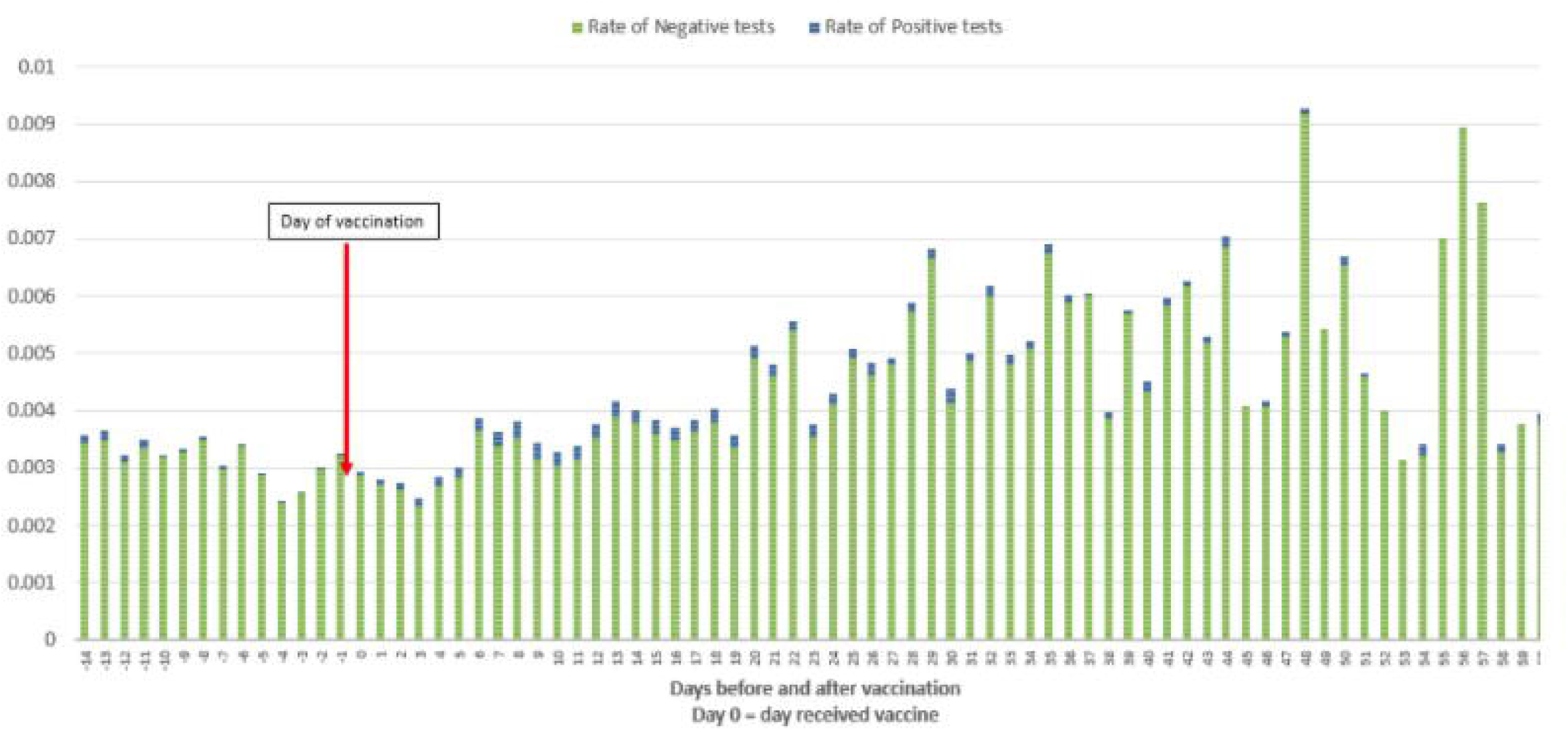
The proportion of all tests (not limited to first positive) per day based on the number of individuals available for follow up split by positive and non-positive results.

In summary, Table 1 shows that infections decrease from day 14 post vaccination to rates that are lower than, or equivalent to, the population weekly levels (Figure 1) and these decreases are not a result of a reduction in testing post vaccination (Figure 4). The risk of COVID 19 infection rate was lower in the vaccinated population than the unvaccinated population (Figure 5). The time to testing positive in the vaccinated group compared with the unvaccinated group was similar until day 15 post vaccination when the groups appear to diverge (Figure 5).

**Figure 5:**
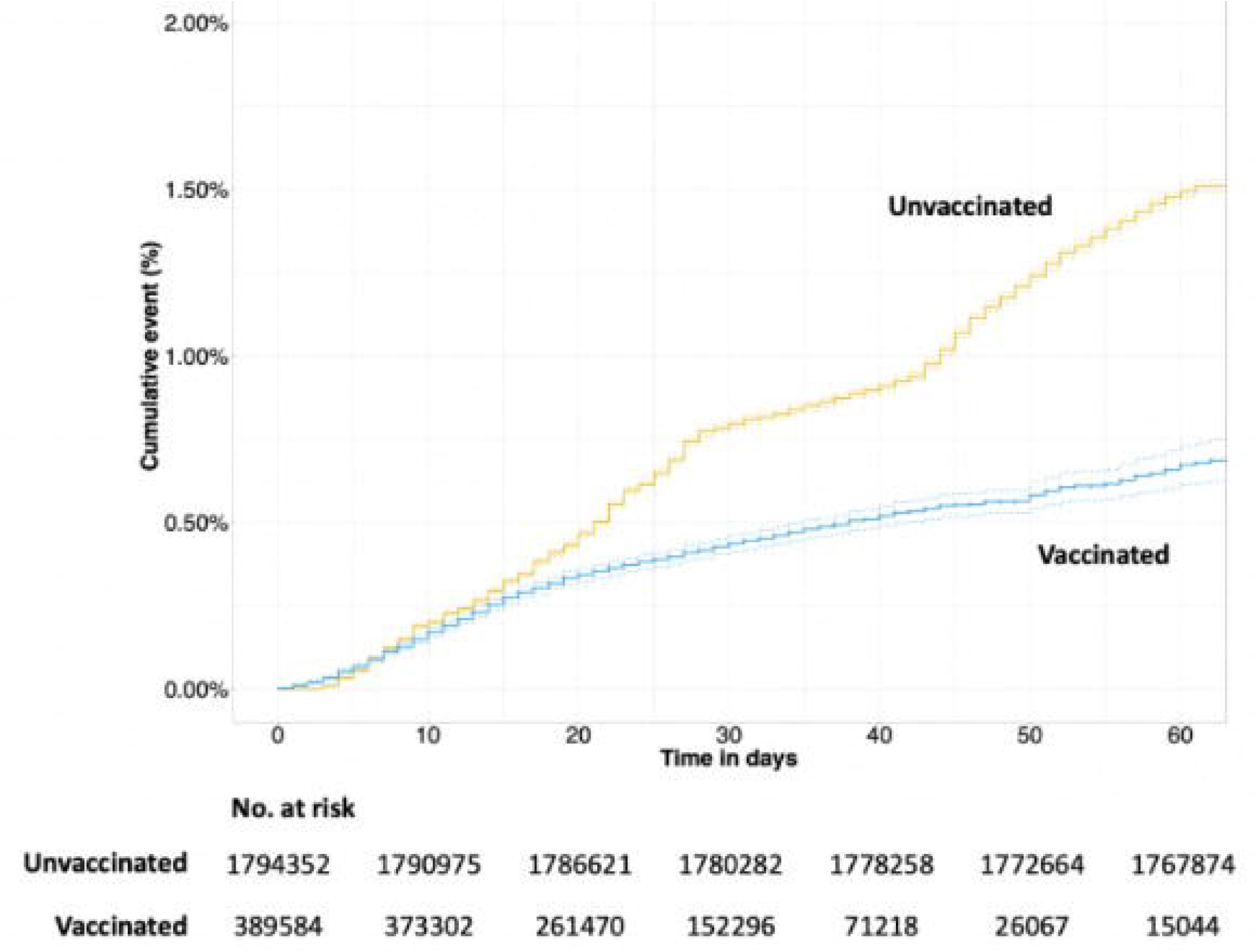
Cumulative event rate with a positive test result in the vaccinated and unvaccinated groups. Numbers at risk are calculated at 10-day intervals. Dotted lines depict 95% confidence intervals.

### Covid-19 vaccination effectiveness

Vaccination effectiveness was measured according to the rates and hazard ratios of testing positive post vaccination compared to the unvaccinated population. In individuals that tested positive post vaccination levels of hospital admissions due to COVID were measured.

Of the eligible vaccination cohort, the average length of follow-up post vaccination was 29 days, with a range of follow-up being 10-79 days. The time to testing positive in the vaccinated group compared with the unvaccinated groups was similar until day 15 post vaccination when the groups appear to diverge, with a smaller cumulative risk in the vaccinated population of testing positive over time (Figure 5).

At 28 days post vaccination there was a 74% (HR 0.26 (0.19-0.35)) and 78% (HR 0.22 (0.18-0.27)) reduction in risk of testing positive for COVID-19 for individuals that received the Oxford/Astrazeneca and Pfizer/BioNTech vaccines respectively, when compared with unvaccinated individuals. There was a lack of significant follow up data in the Oxford/Astrazeneca group to make a meaningful comparison past 28 days, therefore these results are not displayed with HR in table 2. As a reflection of differences in availability of each of the vaccines, patients administered the Pfizer vaccination had longer follow-up to those administered the AstraZeneca vaccine (Figure 4). Accounting for censoring, there were no differences in covid positive event rates comparing people who had the Pfizer and Oxford/AstraZeneca vaccinations (Figure 6).

**Table 2:** Time-dependent Cox Regression Analysis of vaccination effect of COVID-19 positivity during follow-up in all individuals up to 28 days post vaccination.

| Week period | No Vaccination | Oxford/Astrazeneca |  | Pfizer/BioNTech |  |
| --- | --- | --- | --- | --- | --- |
|  |  | HR (95%CI) | P-value | HR (95%CI) | P-value |
| 0 to 7 | 1.0 (Reference) | 0.71<br>(0.60-0.84) | <0.0001 | 1.03<br>(0.91-1.17) | 0.65 |
| 8 to 14 | 1.0 (Reference) | 0.68<br>(0.59-0.80) | <0.0001 | 0.90<br>(0.80-1.00) | 0.06 |
| 15 to 21 | 1.0 (Reference) | 0.59<br>(0.49-0.71) | <0.0001 | 0.42<br>(0.36-0.50) | <0.0001 |
| 22 to 28 | 1.0 (Reference) | 0.26<br>(0.19-0.35) | <0.0001 | 0.22<br>(0.18-0.27) | <0.0001 |

**Figure 6:**
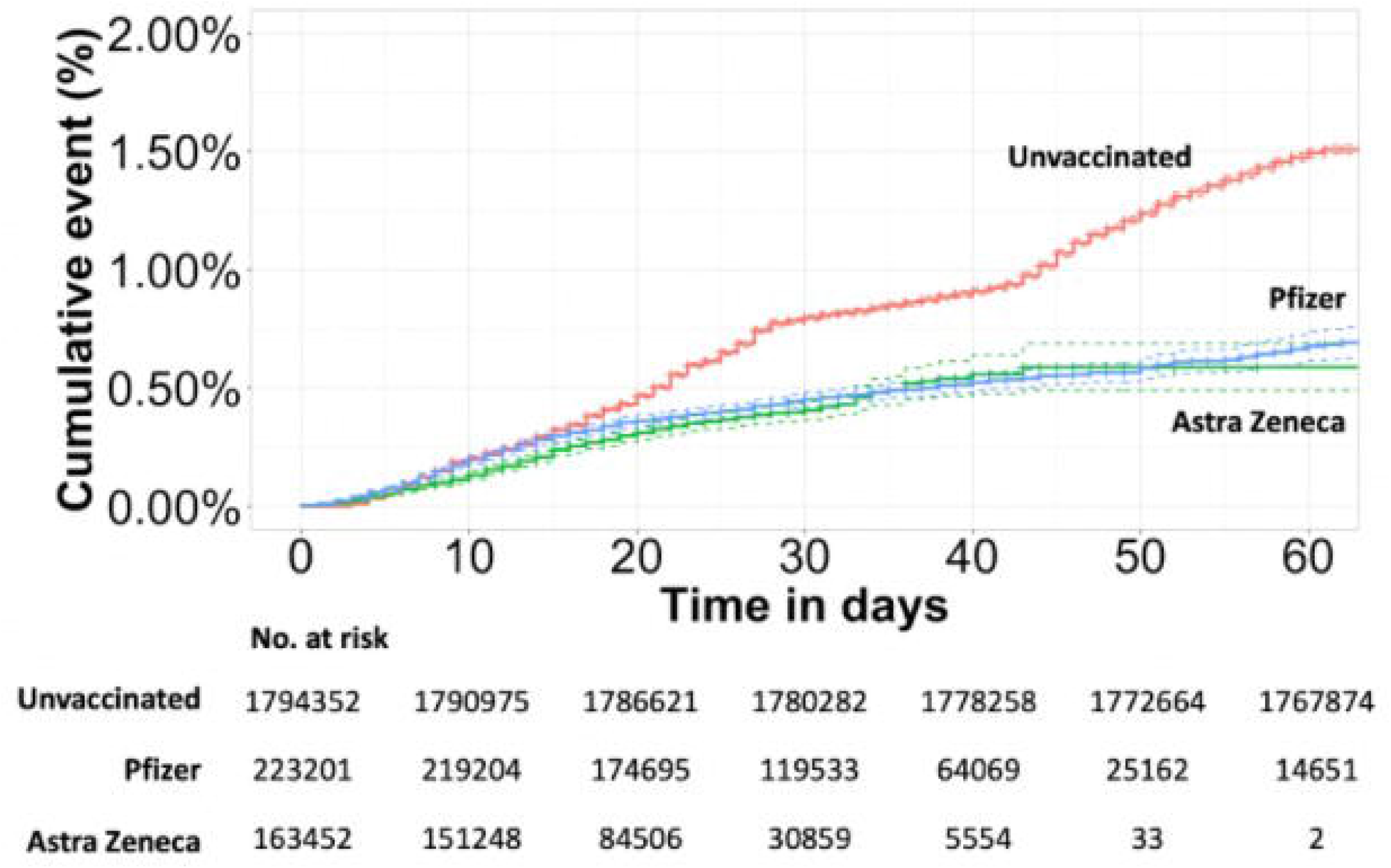
Cumulative event rate of testing positive comparing Pfizer and AstraZeneca vaccination groups to the unvaccinated group. Vaccination type was not available for 2934 patients. Dotted lines depict 95% confidence intervals.

Unvaccinated care home residents were four times as likely compared with individuals aged 16-49 to test positive (HR 4.05 95% CI 3.48-4.71). Unvaccinated Asian or British Asian individuals had a multivariable adjusted hazard ratio (HR) of 1.45 (95% CI 1.41-1.49) of testing positive by 60 days compared to the White group (supplementary Table). All ethnic groups benefited from vaccination with greatest reduction in risk due to vaccination seen in Asian or Asian British individuals (Figure 7). Unvaccinated men were less likely to test positive within 60 days than women (HR 0.89 (95% CI 0.86-0.91)) (supplementary Table), however there was no significant difference between the genders in the vaccinated population (Table 3). There were no significant differences in hazard ratios associated with a positive result with vaccination across ethnicities, IMD decile groups or gender. Significant differences in hazard ratios show that infections in older age groups (65-69, 70-74, 75-80 and 80 +) and in clinically extremely vulnerable were present, showing these groups are significantly less likely to be infected post vaccination, indicating vaccine effectiveness in the oldest population groups (Table 3).

**Table 3:** Multivariable cox regression analysis showing hazard ratio of a positive result during follow-up with vaccination in all patients and across different age, ethnic, gender and IMD decile groups. Cox regression model included an interaction term between having the vaccination and individual patient groups (age, ethnicity, gender, IMD decile).

|  | HR (95% CI) | P-value |
| --- | --- | --- |
| Vaccination |  |  |
| No vaccination (reference) | 1 | - |
| Vaccination | 0.64 (0.43 – 0.95) | 0.03 |
| Age |  |  |
| 16-49 years (reference) | 1 | - |
| 50-54 years | 0.75 (0.58 – 0.98) | 0.03 |
| 55-59 years | 0.82 (0.64 – 1.06) | 0.13 |
| 60-64 years | 0.79 (0.61 – 1.02) | 0.07 |
| 65-69 years | 0.41 (0.32 – 0.54) | <0.0001 |
| 70-74 years | 0.26 (0.20 – 0.33) | <0.0001 |
| 75-79 years | 0.29 (0.23 – 0.38) | <0.0001 |
| 80+ years | 0.29 (0.24 – 0.36) | <0.0001 |
| Care Home Resident | 0.76 (0.56 – 1.05) | 0.1 |
| Clinically extremely vulnerable | 0.30 (0.24 – 0.38) | <0.0001 |
| Ethnicity |  |  |
| White (reference) | 1 | - |
| Asian or asian british | 0.91 (0.80 – 1.02) | 0.11 |
| Black or black british | 0.98 (0.77 – 1.25) | 0.89 |
| Mixed | 1.29 (0.91 – 1.82) | 0.15 |
| Other ethnic groups | 1.06 (0.83 – 1.35) | 0.65 |
| Gender |  |  |
| Female (reference) | 1 | - |
| Male | 1.02 (0.91 – 1.15) | 0.69 |
| IMD decile |  |  |
| 1 (reference) | 1 | - |
| 2 | 1.03 (0.74 – 1.43) | 0.87 |
| 3 | 1.11 (0.82 – 1.51) | 0.49 |
| 4 | 0.97 (0.71 – 1.32) | 0.83 |
| 5 | 1.08 (0.79 – 1.48) | 0.62 |
| 6 | 1.04 (0.76 – 1.42) | 0.79 |
| 7 | 0.80 (0.58 – 1.12) | 0.2 |
| 8 | 0.84 (0.59 – 1.20) | 0.33 |
| 9 | 0.93 (0.65 – 1.34) | 0.7 |
| 10 | 0.83 (0.56 – 1.21) | 0.33 |

**Figure 7:**
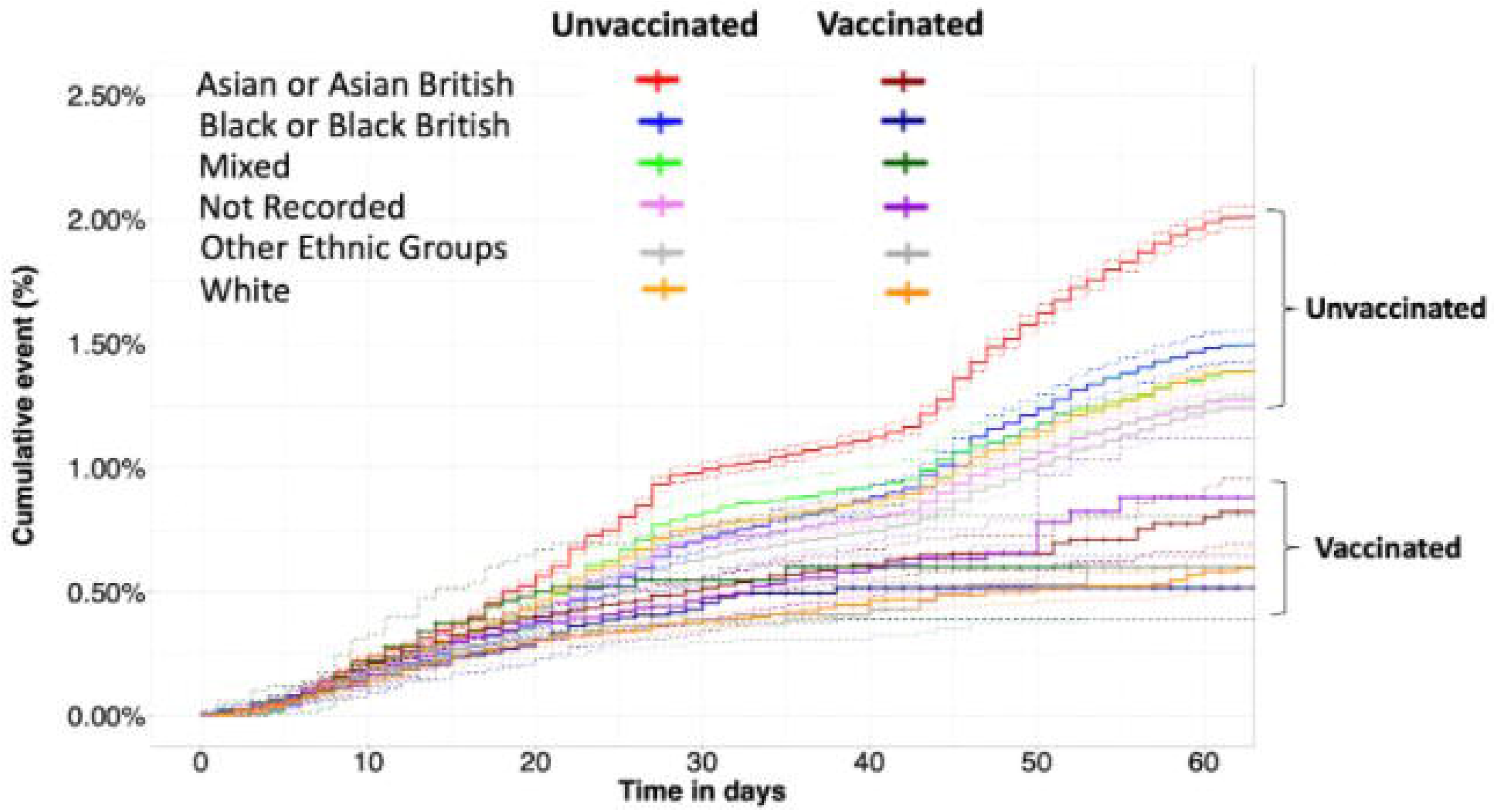
Cumulative event rate of testing positive in the vaccinated and unvaccinated groups stratified by ethnicity. Dotted lines depict 95% confidence intervals.

After vaccination very low rates of hospital admission were seen. In total 288 vaccinated individuals were admitted to hospital post vaccination and tested positive for COVID19 after vaccination and up to 7 days into their admission. Of these patients 54% (155) were admitted before day 14 after vaccination. Admission rates based on individuals available to follow up peaked at 0.026% in days 7-13 and reduced to 0.01% or lower from days 28-34.

Between 8th December 2020 and 24th February 2021, there were a total of 441 all-cause deaths which comprised 161 (36.5%) and 280 (64.5%) in the vaccinated and unvaccinated groups, respectively. Of the 161 deaths in the vaccinated group, 18 (11.2%) had a positive COVID-19 test in the 28 days preceding death (1 in 21,739 of all vaccinated patients). Of the 280 deaths in the unvaccinated group, 68 (24.3%) had a positive COVID-19 test in the 28 days preceding death (1 in 556 of all unvaccinated patients).

## Discussion

By 15th February 2021, the NWL Vaccination programme had vaccinated 17.84% of the eligible population, according to priority, with at least one dose of a COVID-19 vaccine over a 10-week period, commencing 8th December 2020. Understanding and addressing vaccine hesitancy, across the population offered a vaccine, represents an important improvement opportunity to maximise widespread population vaccination coverage; in this study 5.88% of the NWL eligible population declined a vaccine. Rates of vaccine decline within Black and Black British groups were three times greater (16.14%) than the White population. A quarter of Black and Black British individuals who were aged 80+ or clinically extremely vulnerable (27.58% and 23.97%) declined the vaccine. This finding is supported by similar reports examining vaccine hesitancy (13). There was a strong negative correlation between deprivation score and vaccine hesitancy; individuals in the most deprived areas declined vaccinations at a rate 13 times higher than those in the most affluent areas. Overall, across NWL London the highest rates of vaccine decline were seen in elderly and black British people living in the most deprived areas. The causes for this were not assessed by this study but highlights an important area of focus for quality improvement, public and societal engagement and outreach initiatives to improve vaccination coverage across all population groups, especially in relation to findings that indicate vaccine effectiveness.

As previous studies have shown this data supports the strategy of prioritising the elderly and care home residents, as unvaccinated care home residents were four times as likely to test positive (HR 4.05 (CI 3.48-4.71)) compared with individuals aged 16-49. There is further evidence of differing susceptibility to COVID-19 across socio-demographic groups which could support further vaccine prioritisation to those who would benefit most; unvaccinated Asian and Asian British individuals were at increased risk of testing positive for COVID-19 compared to the white population (HR 1.45 (CI 1.41-1.49)) and unvaccinated women more likely to test positive in 60 days than men (male HR 0.89 (CI 0.86-0.91)).

The incubation period to develop symptoms indicative of COVID-19 is on average 5-6 days but can be as long as 14 days (5,7). This means that the majority of transmission at the point of vaccination should be detected and confirmed by positive test results within 14 days of vaccination. The rate of positive COVID-19 cases in the second week (days 7-13) after receiving a vaccine at a vaccination hub, or via a roving team for care home and housebound individuals, peaked at 0.13%. Although this was higher than 0.09% recorded in days 1-6, it was lower than the average weekly person testing positive rate recorded in the total population at 0.19%. This supports the conclusion that the act of vaccine delivery in NWL did not increase COVID-19 virus transmission above that already seen in the background population. Despite overall low levels of positive testing in the vaccinated group however, the increase in positive tests recorded in days 7-13 after vaccination do suggest some potential for increased COVID-19 transmission at or after the time of vaccination. It is impossible to identify and separate out several possible contributors to this, including in the days post-vaccination individuals were more liberal with isolation/social distancing measures before immunity resulting from vaccination had become effective; some transmission of COVID-19 occurring at time of vaccine administration; and/or individuals were asymptomatic but infected when attending for vaccination. Certainly, regarding the latter, there is some evidence to support this as a number of individuals tested positive within 5 days of attending for vaccination (Figure 4).

In the care home residents and/or housebound individuals the rise in positive case rate in the second week post vaccination was greater than that of the rest of the vaccinated population, 0.35% compared to 0.13% in non-care home/housebound individuals. This higher rate needs to be interpreted within the context of physically frail groups having innate vulnerability to COVID-19 transmission (14). Equally, it is not possible to determine the contribution of post-vaccination easing of social distancing and isolation measures prior to the vaccination generating an immune response that provides effective protection. There is also some evidence that the time for the more elderly to develop effective immunity takes longer than the younger population (15). This is supported by a trend suggesting the rate of positivity decreases week on week more slowly when compared with the non-care home/housebound group, but absolute numbers of positive cases in care homes/housebound individuals were very low. These results highlight the importance of maintaining physical COVID-19 restriction procedures post vaccination, particularly in the first fortnight. Care home residents and housebound individuals may be particularly vulnerable in the immediate period post vaccination. Thus, emphasising the need to maintain social distancing and restricting visitors to care homes to prevent exposure until population prevalence of COVID-19 has fallen to sufficient levels to make transmission unlikely and time has elapsed to allow post-vaccination immunity to develop in this higher risk population.

The rise in positive case rates seen in the care home population after the 7th week post vaccination (0.28%) raises concerns that the immunological effects of the single vaccine dose may be waning in the frail elderly population over time, which could be due to immunosenescence. The significance of this, however, needs to be interpreted within the very small numbers completing follow-up in this group (N=1,090). Further studies to examine this are required as it will have implications for timing of second vaccine administration, which may well vary across priority groups.

Overall, in the NWL population, the rate of positive testing in the vaccinated group compared with the unvaccinated group was similar until day 15, whereafter vaccination reduced an individual’s chance of testing positive for COVID-19 beyond 10 weeks of follow-up. The cumulative risk reduction of testing positive for COVID-19 at 60 days was 36% (HR 0.64 (CI 0.43-0.95 p=0.0272)) when receiving a single dose of any vaccine. By the 4th week of follow up (days 22-28) there was similar efficacy for vaccination, with a 74% (HR 0.26 (0.19-0.35)) and 78% (HR 0.22 (0.18-0.27)) reduction in risk of testing positive for COVID-19 in the Oxford/Astrazeneca group and Pfizer/BioNTech group respectively compared with the unvaccinated population. There were insufficient numbers of individuals with enough follow up data in the Oxford/Astrazeneca group to power a statistical comparison between vaccine types beyond 28 days.

The reduction in severity of cases is also evident as demonstrated by the low numbers of admissions to hospitals for vaccinated individuals, with admission rates dropping post 14 days of vaccination. Further work is required to compare admissions in the vaccinated population and comparable control populations, including for non-Covid reasons. The vaccinated and unvaccinated populations are inherently different as vaccination was rolled out according to the priority groups first.

A reduction in the risk of testing positive became apparent from day 15 after the administration of a single dose of vaccine in our study. This finding is similar to phase three trial (3) data showing a benefit from day 10-13 post a first dose in the Pfizer vaccine, and from day 18 in a real-world data study (16).

Interim analysis of four randomised controlled trials in Brazil, South Africa, and the UK examining the safety and the efficacy of the Oxford/Astrazeneca vaccine did not report efficacy data of a first dose before day 21 post vaccination, showing an efficacy of 64·1% (95% CI 50·5–73·9) after 21 days (4). Our study demonstrated an observable reduction in risk of testing positive before 21 days, with a 29% (CI 16 - 40% P<0.001) and 32% (CI 20-41% P<0.001) reduction in the first (days 0-6) and second (days 7-13) week, respectively, after receiving a first dose of Oxford/Astrazeneca (supplementary Figure).

Our findings show at 22-28 days post vaccination there is 78% (HR 0.22 (0.18-0.27)) reduction in risk of testing positive for COVID-19 after a single dose of the Pfizer/BioNTech vaccine in a cohort representative of a UK urban population. This is comparable to real world evidence in an Israeli population administered the Pfizer/BioNTech vaccine showing the early effectiveness of a single dose was estimated to be 52% during the first 24 days after vaccination (16), although a reanalysis of the same data by Hunter et al. estimated that by day 24, vaccine effectiveness had reached 90%.(17) The variation in study design may explain differences seen in efficacy as the Israeli study used the vaccinated population in days 1-12 of vaccination as the control group compared to an unvaccinated control group in our study.

Hall et al. studied the outcomes of the Pfizer/BioNtech vaccine on a cohort of NHS healthcare workers as part of the SIREN study with a similar length of follow up. This prospective cohort study found reduced levels of vaccine coverage in minority groups, especially Black or Black British groups, similar to our findings, even within a healthcare worker population. A single dose of Pfizer/BioNtech vaccine demonstrated vaccine effectiveness of 72% (95% CI 58-86) 21 days after the first dose comparable to our findings although in a purely working age population (18).

Bernal et al. are awaiting peer review of their test negative case control study estimating the effect of vaccination with the BNT162b2 and ChAdOx1 COVID-19 vaccines in an older population of 70 years or older in England. In individuals aged 80+ immunised with BNT162b2 vaccine demonstrated an effectiveness of 70% (95% CI 59-78%) from 28-34 days. ChAdOx1 vaccine effects were seen from 14-20 days after vaccination reaching an effectiveness of 60% (95%CI 41-73%) from 28-34 days and further increasing to 73% (95%CI 27-90%) from day 35 onwards. Similar to our findings Bernal et al. demonstrated an increased risk of testing positive in the first 14 days after receiving a vaccine, and those aged 80+ at particular risk in the first 9 days after vaccination (odds ratio up to 1.48, 95%CI 1.23-1.77) (19).

This study uses a unique linked dataset which provides real time data for clinical and operational care delivery, especially relevant during the COVID-19 pandemic. This study highlights the use of these data for generating real-world evidence in accordance with translational data analytics, in addition to data collected through prospective clinical trials. The large sample size of over 2 million people receiving 389,587 doses of a vaccine is a strength of the study with a comparatively long follow-up time compared to other studies that have been reported to date. The cost of running an RCT of this size would be significant, but equally outcome measurements from real-world evidence are less robust and the results must be interpreted accordingly. The lack of robust control groups to compare with the vaccinated population is problematic, but further analysis similar to methods used by Kaura et al (20) on emulating clinical trials using observational data may be able to address these issues.

The low specificity and sensitivity of some testing mechanisms may provide a degree of error as rates of positive COVID-19 tests are used to estimate COVID-19 prevalence in the population. Test results available included pillar one and two, but not lateral flow test results.

No data were collected on COVID-19 symptoms and so no assessment on the effects of vaccination on COVID-19 symptoms could be made. By capturing only pillars 1 and 2 of testing data this study likely misses asymptomatic cases of COVID-19 in the population, underestimating its true rate. Variation in the prevalence of COVID-19 in the population during the study period could impact the results of the study. Declining rates of COVID-19 in the population during the time of maximal vaccine delivery could have amplified the observed effects of the vaccine.

The likely dominant COVID-19 variant in the examined population at time of study was B1.1.7 (21). Data on COVID-19 variants were not collected during the study. The study findings may not be comparable in populations with differing dominant COVID-19 strains.

Only COVID positive results in the vaccinated group were included in this analysis, therefore we were not able to assess the impact of antibodies developed from previous infection compared with antibodies developed as a result of vaccination. However, there remain other multiple confounders which cannot be determined from the data, namely unconfirmed infections, asymptomatic positive individuals, and the uncertain length of time that post-vaccination immunity persists.

## Conclusion

This study provides further evidence that a single dose of either the Pfizer/BioNTech vaccine or the Oxford/Astrazeneca vaccine is effective at reducing the risk of testing positive for COVID-19 up to 60 days across all adult age groups, ethnic groups, and risk categories in an urban UK population. There was no difference in effectiveness up to 28 days between the Oxford/Astrazeneca and Pfizer/BioNtech vaccines.

By the 15th February 2021, 17.84% of the population in NWL had been vaccinated.In those declining vaccination higher rates were seen in those living in the most deprived areas and in Black and Black British groups.

There was no definitive evidence to suggest COVID-19 was transmitted as a result of vaccination hubs during vaccine the administration roll-out in NWL, and the risk of contracting COVID-19 and/or becoming hospitalised after vaccination has been demonstrated to be very low in the vaccinated population. Individuals appear to be less susceptible to COVID-19 transmission in the first weeks after receiving a vaccine as compared with the unvaccinated population, however, a clear message reinforcing the need to continue social distancing restrictions post vaccination should be delivered at the time of vaccination and potentially for up to 21 days. There is also evidence to suggest that in the care home and housebound population the period of social distancing measures should be more carefully adhered to post vaccination, as initial evidence suggests the time to potentially acquire immunity in this group could take longer than in the general population.

**What is already known on this topic**

⍰ Anticipated vaccination coverage of priority groups has been reduced by vaccine hesitancy, with the use of vaccination centres reported as being a potential contributory factor.
⍰ There is no real-world evidence, to date, to understand the rates of COVID-19 positive testing after vaccination.
⍰ There is little evidence to assess the early effectiveness of Covid-19 vaccination stratified across population sub-groups and by vaccine type and compared with rates of COVID-19 positive testing rates in the non-vaccinated population.

**What this study adds**

⍰ Rates of declining a vaccine varied considerably across ethnic groups and when stratified by age. There was also a strong negative correlation between socio-economic deprivation and vaccine uptake.
⍰ The time to testing positive in the vaccinated group compared with the unvaccinated group was similar until day 15 post vaccination when the groups appear to diverge, thus highlighting the importance of maintaining physical COVID-19 restriction procedures post vaccination, particularly in the first fortnight. Care home residents and housebound individuals may be particularly vulnerable in the immediate period post vaccination.
⍰ A single dose of either the Pfizer/BioNTech vaccine or the Oxford/Astrazeneca vaccine is effective at reducing the risk of testing positive for COVID-19 up to 60 days across all adult age groups, ethnic groups, and risk categories in an urban UK population.

## Data Availability

Data sharing

We are unable to extract or publish patient level data from iCARE and Whole systems integrated care due to data protection restrictions. Any request to access data can be made to referring to the title of this paper.

## Acknowledgements

The authors would like to acknowledge members of the NWL Data & Analytics Gold Command for contribution to interpretation of the findings; Roger Chinn, Martin Kuper, Jacques Naude, Merav Dover, James Biggin-Lamming, Sanjay Gautama, Kevin Jarrold. The authors would also like to acknowledge Owain Griffiths from Whole Systems Integrated Care for helping with Data access and data quality checks.

This work uses data provided by patients and collected by the NHS as part of their care and support. Using patient data is vital to improve health and care for everyone. There is huge potential to make better use of information from people’s patient records, to understand more about disease, develop new treatments, monitor safety, and plan NHS services. Patient data should be kept safe and secure, to protect everyone’s privacy, and it’s important that there are safeguards to make sure that it is stored and used responsibly. Everyone should be able to find out about how patient data is used. #datasaveslives

## Contributors

BG, JB, JR, AK, IG and EKM conceived the study aims and objectives. AM and LM carried out the programming to extract and curate the data from the source data tables. BG, AK and EKM undertook all data analyses. JB, BG, AK and EKM drafted the manuscript. BG, JB, AK, AM, LM, SB, PA, TS, IG, JR, KS and EKM provided a critical review of the manuscript. All authors read and approved the final version of the manuscript. EKM is guarantor for this paper. The corresponding author attests that all listed authors meet authorship criteria and that no others meeting the criteria have been omitted.

## Copyright/license for publication

The Corresponding Author has the right to grant on behalf of all authors and does grant on behalf of all authors, a worldwide licence to the Publishers and its licensees in perpetuity, in all forms, formats and media (whether known now or created in the future), to i) publish, reproduce, distribute, display and store the Contribution, ii) translate the Contribution into other languages, create adaptations, reprints, include within collections and create summaries, extracts and/or, abstracts of the Contribution, iii) create any other derivative work(s) based on the Contribution, iv) to exploit all subsidiary rights in the Contribution, v) the inclusion of electronic links from the Contribution to third party material where-ever it may be located; and, vi) licence any third party to do any or all of the above.

## Data sharing

We are unable to extract or publish patient level data from the iCARE and Whole systems integrated care due to data protection restrictions. Any request to access data can be made to referring to the title of this paper.

## Transparency statement

The guarantor (EKM) affirms that the manuscript is an honest, accurate, and transparent account of the study being reported; that no important aspects of the study have been omitted; and that any discrepancies from the study as originally planned (and, if relevant, registered) have been explained.

## Figure Legends

**Supplementary Figure 1:**
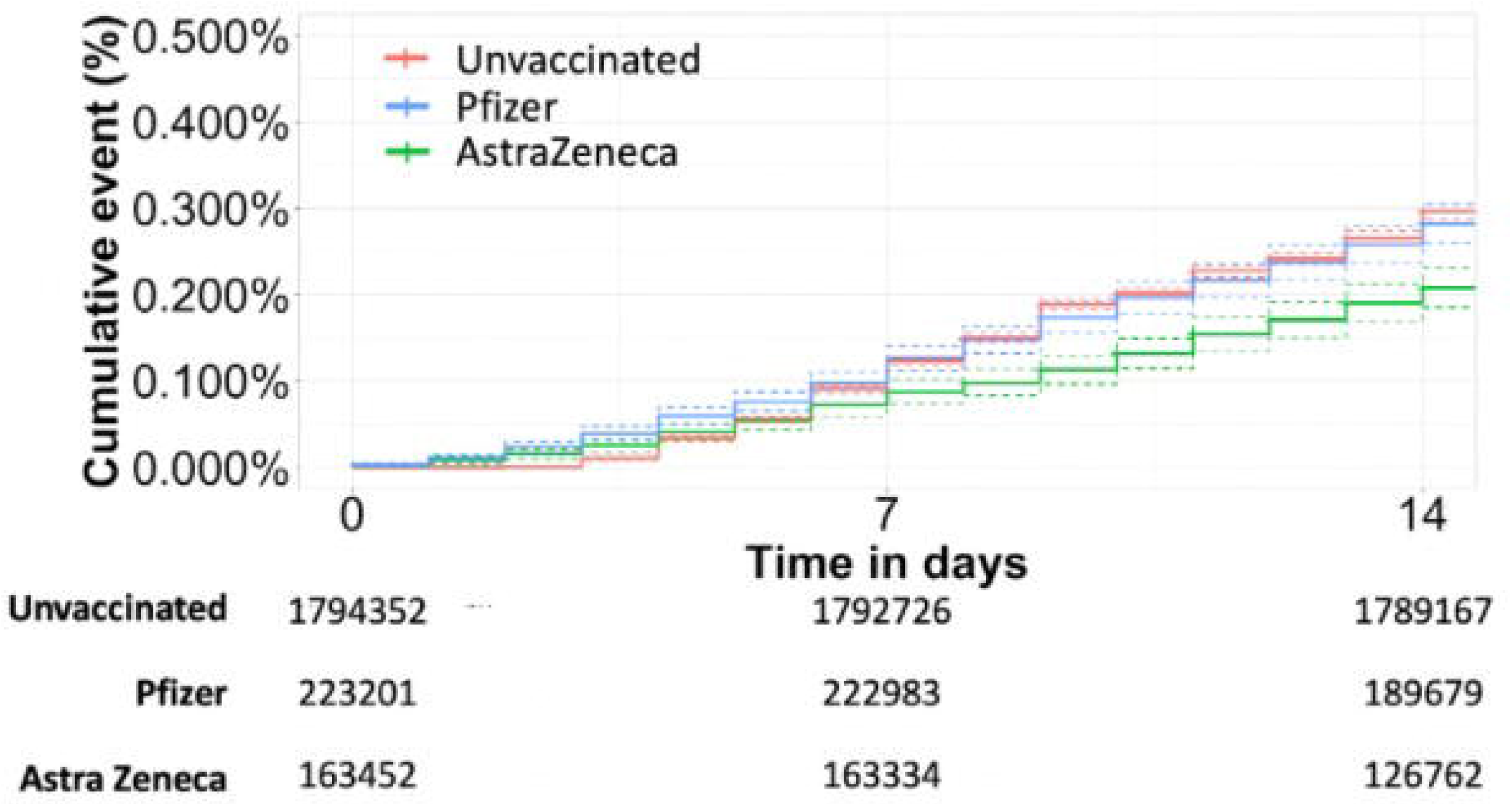
Cumulative event rate of testing positive over the first 2 weeks comparing Pfizer and AstraZeneca vaccination groups to the unvaccinated group. Dotted lines depict 95% confidence intervals

## Table Titles

**Supplementary Table 1:**
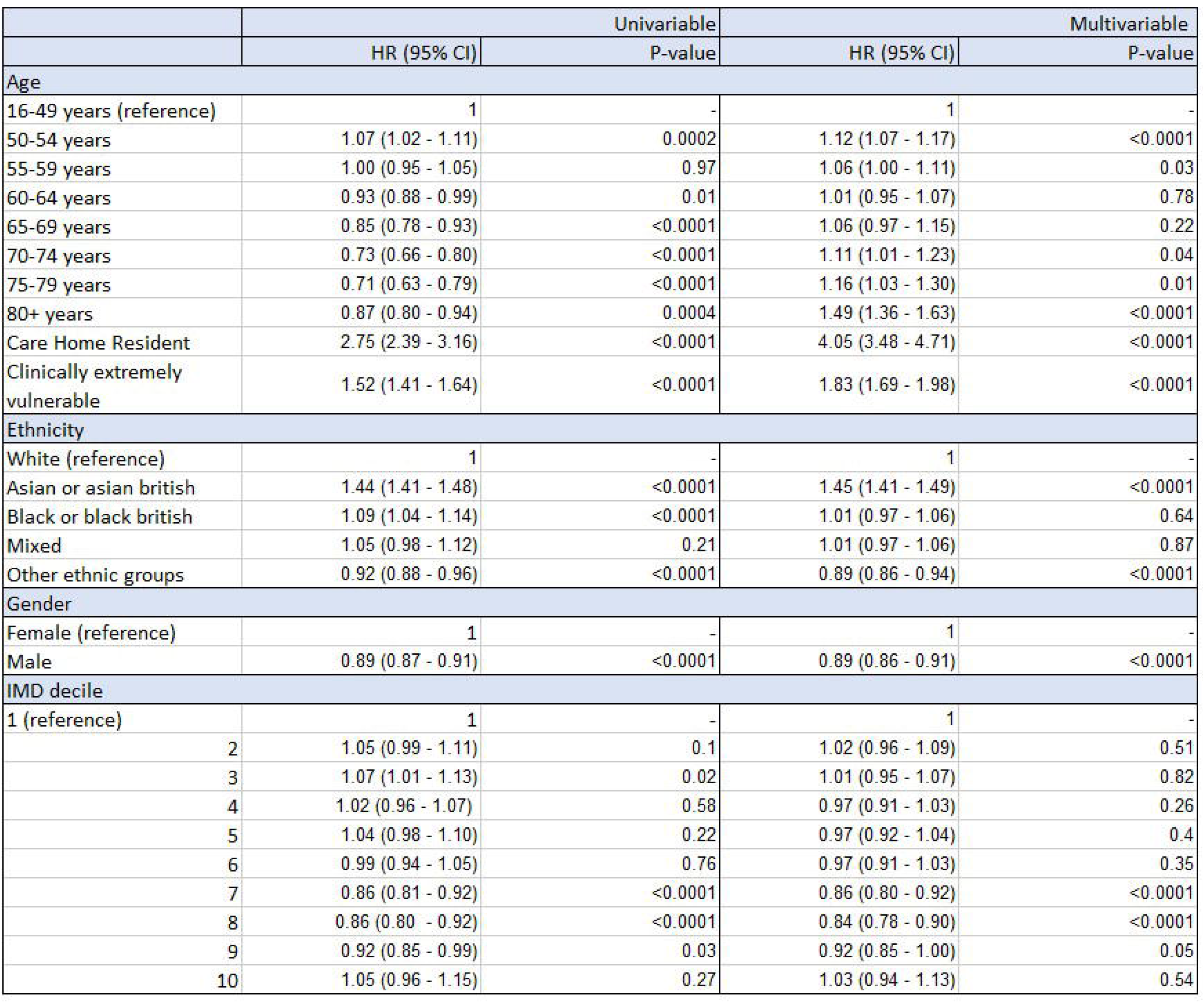
Multivariable Cox regression analysis showing hazard ratio of a positive result in the unvaccinated group across different age, ethnic, gender and IMD decile groups.

|  | Univariable |  | Multivariable |  |
| --- | --- | --- | --- | --- |
|  | HR (95% CI) | P-value | HR (95% CI) | P-value |
| Age |  |  |  |  |
| 16-49 years (reference) | 1 | - | 1 | - |
| 50-54 years | 1.07 (1.02 - 1.11) | 0.0002 | 1.12 (1.07 - 1.17) | <0.0001 |
| 55-59 years | 1.00 (0.95 - 1.05) | 0.97 | 1.06 (1.00 - 1.11) | 0.03 |
| 60-64 years | 0.93 (0.88 - 0.99) | 0.01 | 1.01 (0.95 - 1.07) | 0.78 |
| 65-69 years | 0.85 (0.78 - 0.93) | <0.0001 | 1.06 (0.97 - 1.15) | 0.22 |
| 70-74 years | 0.73 (0.66 - 0.80) | <0.0001 | 1.11 (1.01 - 1.23) | 0.04 |
| 75-79 years | 0.71 (0.63 - 0.79) | <0.0001 | 1.16 (1.03 - 1.30) | 0.01 |
| 80+ years | 0.87 (0.80 - 0.94) | 0.0004 | 1.49 (1.36 - 1.63) | <0.0001 |
| Care Home Resident | 2.75 (2.39 - 3.16) | <0.0001 | 4.05 (3.48 - 4.71) | <0.0001 |
| Clinically extremely vulnerable | 1.52 (1.41 - 1.64) | <0.0001 | 1.83 (1.69 - 1.98) | <0.0001 |
| Ethnicity |  |  |  |  |
| White (reference) | 1 | - | 1 | - |
| Asian or asian british | 1.44 (1.41 - 1.48) | <0.0001 | 1.45 (1.41 - 1.49) | <0.0001 |
| Black or black british | 1.09 (1.04 - 1.14) | <0.0001 | 1.01 (0.97 - 1.06) | 0.64 |
| Mixed | 1.05 (0.98 - 1.12) | 0.21 | 1.01 (0.97 - 1.06) | 0.87 |
| Other ethnic groups | 0.92 (0.88 - 0.96) | <0.0001 | 0.89 (0.86 - 0.94) | <0.0001 |
| Gender |  |  |  |  |
| Female (reference) | 1 | - | 1 | - |
| Male | 0.89 (0.87 - 0.91) | <0.0001 | 0.89 (0.86 - 0.91) | <0.0001 |
| IMD decile |  |  |  |  |
| 1 (reference) | 1 | - | 1 | - |
| 2 | 1.05 (0.99 - 1.11) | 0.1 | 1.02 (0.96 - 1.09) | 0.51 |
| 3 | 1.07 (1.01 - 1.13) | 0.02 | 1.01 (0.95 - 1.07) | 0.82 |
| 4 | 1.02 (0.96 - 1.07) | 0.58 | 0.97 (0.91 - 1.03) | 0.26 |
| 5 | 1.04 (0.98 - 1.10) | 0.22 | 0.97 (0.92 - 1.04) | 0.4 |
| 6 | 0.99 (0.94 - 1.05) | 0.76 | 0.97 (0.91 - 1.03) | 0.35 |
| 7 | 0.86 (0.81 - 0.92) | <0.0001 | 0.86 (0.80 - 0.92) | <0.0001 |
| 8 | 0.86 (0.80 - 0.92) | <0.0001 | 0.84 (0.78 - 0.90) | <0.0001 |
| 9 | 0.92 (0.85 - 0.99) | 0.03 | 0.92 (0.85 - 1.00) | 0.05 |
| 10 | 1.05 (0.96 - 1.15) | 0.27 | 1.03 (0.94 - 1.13) | 0.54 |

